# Investigation of the continuous spread of SARS-CoV-2 in the post pandemic time - Insights into the reason for the sustained spread despite the establishment of population immunity

**DOI:** 10.64898/2026.06.05.26355009

**Authors:** Buqing Yi

## Abstract

In spite of well-established global immune landscape, SARS-CoV-2 is still able to further spread and continue causing infection waves. The current understanding about the reason behind is limited, and it is still difficult to predict the evolution or spreading tread of SARS-CoV-2. Therefore, it is necessary to investigate whether the establishment of population immunity has changed the virus evolution or spreading pattern.

In this investigation, one overall analysis of the SARS-CoV-2 spreading in the past several years have been carried out through one thorough genomic epidemiology study, with Germany being chosen as one representative location in view of the systemic efforts for genomic surveillance. The growth advantage of a few predominant variants in its early spreading period has been evaluated through a logistic regression model. The results have revealed that the major circulating SARS-CoV-2 variants since 2023 are mainly derived from the Omicron BA.2 family. Since middle of 2024, most predominant variants were produced primarily through recombination, indicating that the evolution derived from recombination might be the major driving force for the continuous spread of SARS-CoV-2 despite the existence of population immunity. Furthermore, the lower growth advantage of recently emerged variants might possibly lead to a tread of reduction in the frequency of infection wave.

The information revealed from this investigation suggests that although short-term spreading tread can be affected by specific virus feature as well as local immunity landscape, the long-term spreading tread is mainly decided by the genomic diversity of the viruses, and can be predicted through phylogenetic and genomic epidemiology investigation. The results have emphasized the importance of maintaining the efforts for genomic surveillance of SARS-CoV-2, which is essential from both medical and research perspectives.

## 1. Introduction

Despite the built population immunity against SARS-CoV-2, the virus further spread worldwide, and caused one infection wave after another. What is the reason for the continuous spreading of this virus? Why the population immunity could not stop its spreading? Till now our understanding about this important topic is still limited. And it remains difficult to predict the future tread of spreading and evolution of SARS-CoV-2. One assumption is that the continuous evolution of SARS-CoV-2 might have played one important role here ^1^.

In addition to random mutations, recombination events can also promote the evolution of SARS-CoV-2. It has been reported that recombination between different coronaviruses might have led to the origin of SARS-CoV-2 ^2-5^. Recombinant virus variants contain a unique combination of mutations which have possible influences on virulence or other functional aspects of the virus ^6-8^. It is therefore important to track recombinant genomes. In the first year of the pandemic, certain SARS-CoV-2 recombinants already emerged and circulated ^9^. Following the designation of XA, the first identified SARS-CoV-2 recombinant lineage in the Pango nomenclature system ^10^, till now a large number of recombinant lineages have been identified ^11^, such as the variant XBB, XDE and XEC.

In our recent papers, comprehensive analyses about recombination events of SARS-CoV-2 as well as the evolution of Omicron variant were provided ^12,13^, which have revealed that NB.1.8.1 has the potential to become one predominant variant and one important source for evolution because it combines the genetic feature of two distantly related virus families, manifesting a higher level of genetic divergence from other strains. It is necessary to follow up and investigate, if this prediction reflects the reality. Furthermore, it is a good timing to run one overall analysis about the spread of SARS-CoV-2 in the past several years after the establishment of population immunity, to detect if there are any rules that could help predict the future tread.

In Germany, SARS-CoV-2 surveillance has been carried out systematically, and sequencing efforts have been made to achieve high-quality genomic data ^14,15^. Till now, most designated SARS-CoV-2 lineages and a few newly emerging variants have been identified through the genomic surveillance efforts in Germany ^16-18^. Considering the high-quality data and standardized system, in this paper, we have chosen Germany as one representative location to analyze the virus spreading in the post-pandemic time.

## 2. Methods

### 2.1 Investigating the spread of SARS-CoV-2 in Germany through genomic epidemiology

For genomic epidemiology investigation, we used samples collected in Germany and uploaded to GISAID ^19^. Only samples fulfilling these quality check criteria on GISAID were included in the analysis: 1. With a complete sequence (>29,000nt) and less than 5% Ns; 2. With complete sample collection dates. We then performed quality check and filtered out low-quality sequences that met any of the following criteria: 1) genomes with too many private mutations (defined as having >24 mutations relative to the closest sequence in the reference tree); 2) genomes with mutation clusters, defined as 6 or more private differences within a 100-nucleotide window. These are the standard quality assessment parameters ^20^.

Thereafter, we combined SARS-CoV-2 sequences that passed the quality check to build up genome sequence data set for epidemiology investigation. In the current study, between January 2022 and February 2026, more than 550,000 sequences from Germany were used in the genomic epidemiology analyses. The analyses were based on genome data from GISAID acquired on April 30, 2026.

For lineage identification, we used the dynamic lineage classification method through the Phylogenetic Assignment of Named Global Outbreak Lineages (PANGOLIN) software suite (https://github.com/hCoV-2019/pangolin) ^21^.

### 2.2 Relative growth advantage

We also analyzed relative growth advantage with SARS-CoV-2 sequences generated from samples collected in Germany from the defined time duration through a logistic regression model. As previously reported ^22-25^, the logistic regression model was applied to evaluate the relative growth advantage of certain variant compared to co-circulating variants in the selected region and time frame. The relative growth advantage per week (in percentage; 0% means equal growth) is presented ^26^. The analyses were primarily carried out with RStudio v1.3.1093 with multiple R software, e.g. tidyverse, ggplot ^27-35^.

## 3. Results

### 3.1 SARS-CoV-2 spreading in Germany, from 2022 till early 2026

To acquire detailed information about the spread of SARS-CoV-2 in Germany, we investigated the lineage dynamic changes of SARS-CoV-2 between 2022 and early 2026.

In 2022, the predominant lineages are the earliest Omicron lineages BA.1, BA.2 and BA.5 and their subvariants, labelled as BA.1*, BA.2* and BA.5* in Figure 1. In 2023, the predominant lineages are mostly from the XBB family, such as XBB.1.5* and EG.5.1*. In view of the fact that XBB is recombinant of two BA.2 subvariants ^13,36^, the XBB family was derived from BA.2, instead of BA.5, although in late 2022, BA.5* was the predominant lineage. In late 2023, as one decedent of BA.2.86 ^13,36^, JN.1 occurred and became the predominant lineage, as shown in Figure 1. Across 2024, the predominant lineages were mostly derived from the BA.2.86/JN.1 family, such as KP.3.1* and XEC* (recombinant of two JN.1 subvariants) ^10,12^. In 2025, XEC* was replaced by a few newly emerged recombinant variants, in particular XFG (also recombinant of two JN.1 subvariants) ^10,12^ and NB.1.8.1.

**Figure 1:**
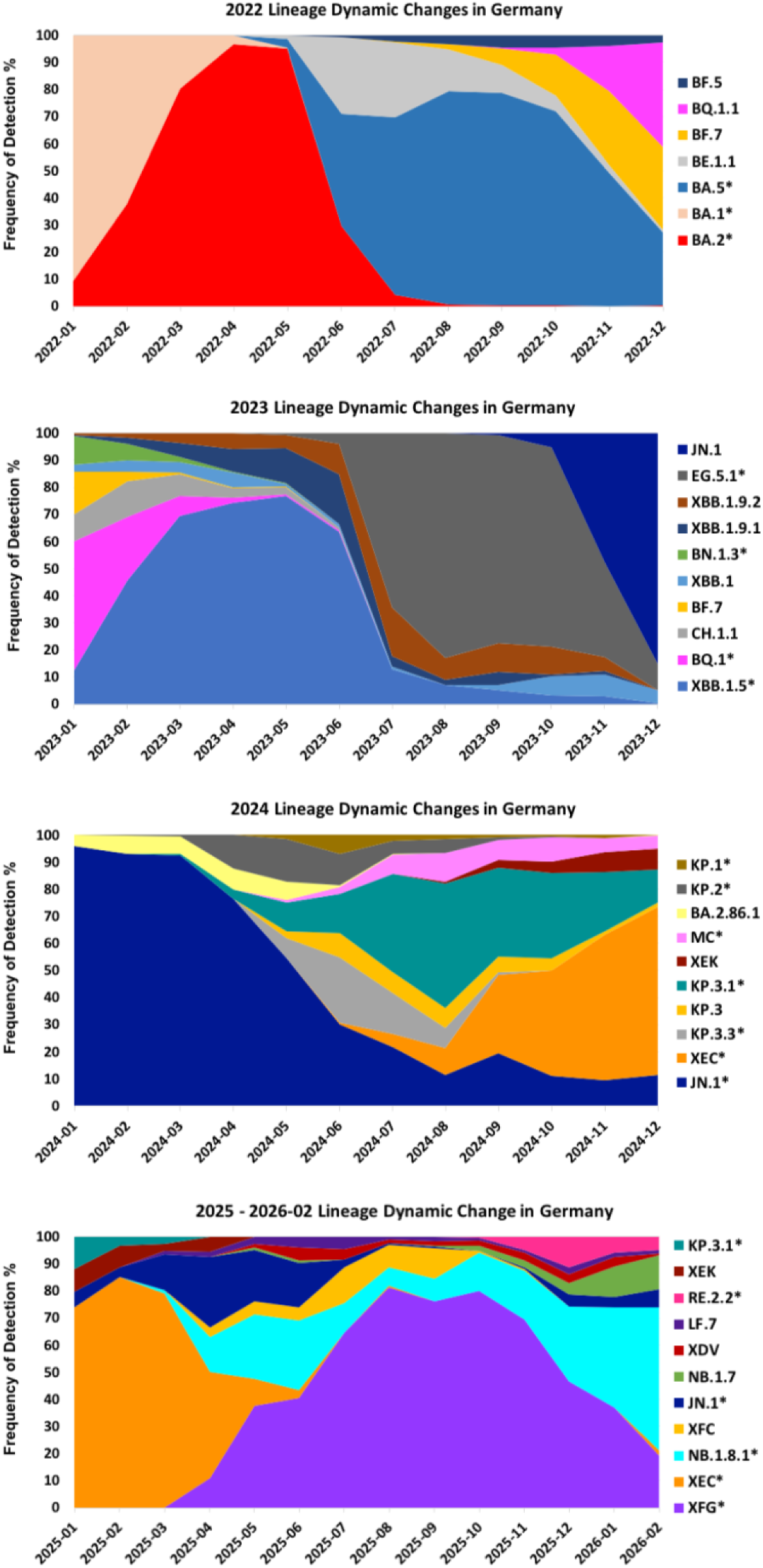
SARS-CoV-2 lineage dynamic changes in Germany, between 2022-01 and 2026-02. Frequency of detection (%) of each SARS-CoV-2 lineage in each month of 2022 (upper), 2023 (upper middle), 2024 (lower middle) and 2025/2026 (lower) is displayed. To achieve a better resolution, if several subvariants of one lineage were detected during this period, in most cases the subvariants are collectively shown together with the ancestor lineage, labelled as the ancestor lineage with star, such as BQ.1* indicating BQ.1 and subvariants. If one specific subvariant is showed separately, the number of this subvariant is excluded from the parent lineage family.

NB.1.8.1 was indirectly derived from recombinant between one XBB subvariant and JN.1^10,37^, showing the highest level of divergence compared to other co-existing variants as being analyzed in one of our recent papers ^13^. Although NB.1.8.1 and XFG have a similar evolutionary distance from the predicted earliest Omicron lineage, in comparison with XFG, NB.1.8.1 is genetically more divergent from other co-existing variants in clustering relationship ^12^.

It is noteworthy that although in the middle of 2025, XFG* surpassed NB.1.8.1* and became the predominant lineage, NB.1.8.1* displayed a growth advantage over XFG* in late 2025 and outcompeted XFG* in early 2026, becoming the predominant lineage, suggesting that the long-term spreading tread is mainly decided by the genetic diversity of the viruses. Also, NB.1.8.1 has further evolved and led to the creation of multiple subvariants, such as PQ.2 and PQ.14, which have also spread widely (In Figure 1, they are included in the NB.1.8.1*).

### 3.2 Relative growth advantage of the predominant variants

Usually the predominant lineages display certain growth advantage over co-circulating variants, and such growth advantage may to certain extent decide the spreading pattern. Therefore, we analyzed the relative growth advantage of several representative predominant lineages during their early spreading period in Germany, namely XBB.1.5, JN.1 and XEC.

The relative growth advantage reflects the advantage over co-circulating lineages in the selected region and time frame. As it has been revealed by previous studies that large scale community transmission often started or became detectable around one month later after the first sample was detected ^36^, to evaluate the growth advantage in the early spreading period, we focus on the time period of about 45 days right after community transmission started.

In Germany, XBB.1.5 showed a relative growth advantage of 60% during its early spreading period between 2023-01-01 and 2023-02-15 (YYYY-MM-DD) over co-circulating variants, mainly BA.5 sub-lineages, such as BF.7, BQ.1 and its subvariants, e.g. BQ.1.1 (Figure 2). In late 2023 between 2023-10-16 and 2023-11-30, JN.1 displayed a strong relative growth advantage of 75% over the co-circulating XBB sub-lineages, after the XBB* family dominating for around 10 months. JN.1 has produced many sub-lineages, some of which were independently identified as VOI/VUM, such as XEC. During its early spreading period in Germany between 2024-08-01 and 2024-09-15, XEC had a relative growth advantage of 38% over co-circulating variants, mainly other JN.1 sub-lineages. Possibly owing to the closely related genetic backgrounds, the relative growth advantage of XEC is lower than that of XBB.1.5 and JN.1 during their own early spreading period. Interestingly, the relative growth advantage of XEC is similar to that of XFG during its early spreading period in Germany (40%) as previously reported ^12^.

**Figure 2:**
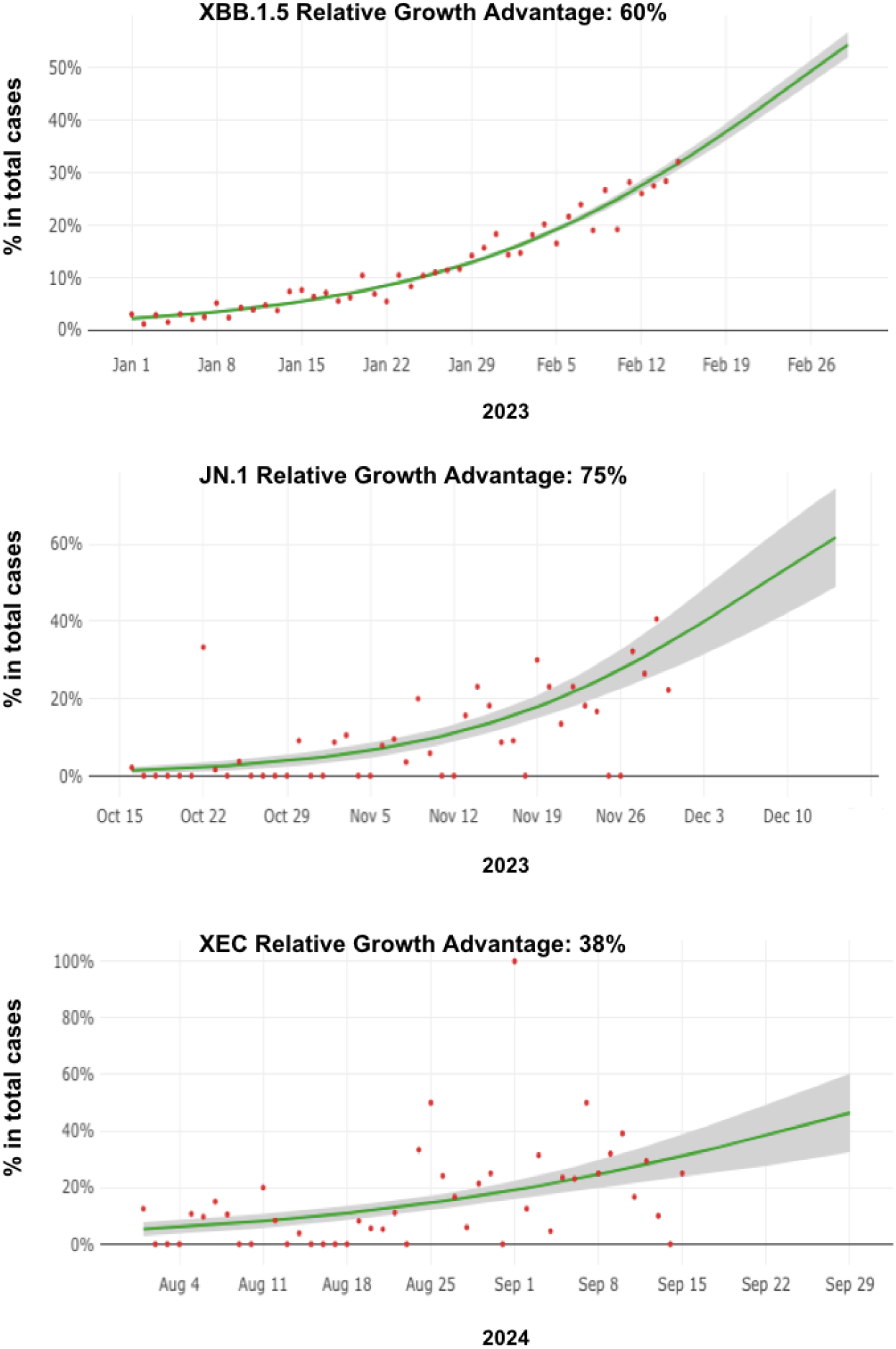
Growth of XBB.1.5, JN.1 and XEC during their early spreading period in Germany. Model fits are based on a logistic regression. Dots represent the daily proportions of variants. The relative growth advantage per week (in percentage; 0% means equal growth) is reported. The shaded areas correspond to the 95% CIs of the model estimates. Relative growth advantage of XBB.1.5 (upper), JN.1 (middle) and XEC (lower) in Germany are displayed.

## 4. Discussion

This investigation has revealed that the major circulating SARS-CoV-2 variants in Germany since 2023 are mainly derived from the Omicron BA.2 family. Since middle of 2024, most predominant variants were produced primarily by recombination, either through recombination events between closely related variants, such as XEC or XFG as recombinant of two BA.2.86/JN.1subvariants, or through recombination events between distantly related variants, such as NB.1.8.1, derived from recombinant of JN.1 and one XBB subvariant. NB.1.8.1* has been spreading steadily since middle of 2025 and outcompeted XFG* in early 2026. The results indicate that the emergence of highly divergent recombinant variants might be the major driving force for the continuous spread of SARS-CoV-2 despite the existence of population immunity.

Recombinant SARS-CoV-2 genomes arise from recombination of two or more different SARS-CoV-2 lineages and often emerge following the co-circulation of multiple lineages at high prevalence, which may lead to co-infection of certain individuals. Co-infection provides the circumstances during which chimeric genotypes can emerge ^38^. Multiple studies have reported identification of recombination events within a single patient co-infected with co-circulating lineages ^39,40^. Nevertheless, there is also genomic evidence indicating that recombination between non-co-circulating viruses, such as one earlier variant with one more recent variant from a subsequent infection, may occur within long-term-infected individual ^40,41^.

Recombinant genomes contain a unique combination of mutations, which could possibly bring certain advantage for the virus ^6,7^. Based on the results of this study, what can be assumed is: there are two major factors affecting the spread of certain virus: 1. Specific virus feature that could bring certain advantage for the virus and consequently affect the short-term spreading. 2. The genetic divergence of one specific virus from others, which could very likely define the base for immune escape capability against general population immunity acquired from previous infection or vaccination, and consequently affect the long-term spreading of this virus. In particular, along with further evolution, certain additional mutant on that virus could increase fitness or other functional features. Consequently, it might be just a matter of time for genetically highly divergent new virus to become the predominant variant.

JN.1 is one example for this scenario. When its ancestor BA.2.86 was first detected in 2023, owing to its relatively low transmission capability, it was assumed that BA.2.86 might not be able to affect the virus spreading tread ^42,43^. But with the emergence of its sub-lineage JN.1 ^36,44^, this changed the spreading pattern dramatically ^45^. Most SARS-CoV-2 variants circulating today are still the decedents of BA.2.86/JN.1^13^. For example, XFG is one sub-lineage of the BA.2.86/JN.1 family ^12^, and NB.1.8.1 also partly carries the genetic background of BA.2.86/JN.1 ^13^. This means, the emergence of the highly divergent variant BA.2.86 has shaped the spreading landscape of SARS-CoV-2 for several years already.

In the current study, it is shown that the genetically more divergent NB.1.8.1 could outcompete other co-existing variants, such as XFG, although XFG is highly competitive with a verified high-level antibody evasion ^46,47^. This finding corresponds to our assumption that the genetic diversity can define the long-term spreading pattern, while specific virus feature may only affect the short-term spreading tread, such as the strong antibody evasion can fade away with the time.

What is the future tread? In view of the fast evolution of the virus, in particular the recombination-derived evolution, the virus might be able to further spread. Taking into account of the built population immunity, generally the emergence of new SARS-CoV-2 variants would not pose a high risk on the public health. Based on the analysis about the growth advantage of newly emerging variants, very likely the chance that one newly emerging variant displays a strong growth advantage might become lower than before in view of the more and more similar genetic background of co-circulating variants, which can lead to a tread of reduction in the frequency of infection wave.

However, this could not exclude the possibility that one newly emerging variant might have higher pathogenicity, such as owing to special structure changes. Owing to the complicated influential factors for pathogenicity, the most effective way to prevent health cost is to keep genomic surveillance to monitor the evolution and spreading constantly.

The information acquired from this study indicates that short-term spreading tread can be affected by specific virus feature as well as local immunity landscape, while long-term spreading tread can be predicted, or at least to a large extent, by investigating genomic diversity and evolution through phylogenetic and genomic epidemiology strategies. To prevent outburst of virus infection case, it is necessary to keep the genomic surveillance of SARS-CoV-2 virus, which is important in view of research purpose as well as public health protection.

## Data Availability

All data produced in the present work are contained in the manuscript

## Acknowledgements

We thank all researchers who generate and share genome data on GISAID (http://www.gisaid.org). We thank the Dresden-concept Genome Centre for their sequencing efforts.

## Code availability

Data processing and visualization was performed using publicly available software, primarily RStudio v1.3.1093.

